# Vaccine-elicited B and T cell immunity to SARS-CoV-2 is impaired in chronic lung disease patients

**DOI:** 10.1101/2023.01.25.23284971

**Authors:** Haolin Liu, Katja Aviszus, Pearlanne Zelarney, Shu-Yi Liao, Anthony N. Gerber, Barry Make, Michael E. Wechsler, Philippa Marrack, R. Lee Reinhardt

**Author notes:** **Material and Correspondence:** Correspondence and material requests should be directed to R. Lee Reinhardt, Haolin Liu, Michael Wechsler, and Phillipa Marrack.

## Abstract

The protection afforded by vaccination against severe acute respiratory syndrome coronavirus 2 (SARS-CoV-2) to individuals with chronic lung disease is not well established. To understand how chronic lung disease impacts SARS-CoV-2 vaccine-elicited immunity we performed deep immunophenotyping of the humoral and cell mediated SARS-CoV-2 vaccine response in an investigative cohort of vaccinated patients with diverse pulmonary conditions including asthma, chronic obstructive pulmonary disease (COPD), and interstitial lung disease (ILD). Compared to healthy controls, 48% of vaccinated patients with chronic lung diseases had reduced antibody titers to the SARS-CoV-2 vaccine antigen as early as 3-4 months after vaccination, correlating with decreased vaccine-specific memory B cells. Vaccine-specific CD4 and CD8 T cells were also significantly reduced in patients with asthma, COPD, and a subset of ILD patients compared to healthy controls. These findings reveal the complex nature of vaccine-elicited immunity in high-risk patients with chronic lung disease.

## Introduction

Vaccination against severe acute respiratory syndrome coronavirus 2 (SARS-CoV-2) targeting the ancestral (Wuhan-Hu-1/2019) viral spike (S) protein has been broadly effective at limiting infection and severe coronavirus disease (COVID-19) (1-6). With respect to SARS-CoV-2 infection, both the humoral and cell mediated arms of the adaptive response are important for achieving optimal control of COVID-19 (7). As such, generating effective B cell and T cell immunity against SARS-CoV-2 remains the goal during vaccination. Much of the protection afforded by both the Pfizer/BioNTech BNT162b2 and the Moderna mRNA-1273 mRNA vaccines is mediated by increased serum neutralizing antibodies to the viral spike protein (8). The efficacy of such neutralizing antibodies depends on their titer, avidity, and half-life (9-17). Indeed, the importance of maintained humoral immunity is evident since breakthrough cases of COVID-19 appear in otherwise healthy, vaccinated or previously infected individuals at the time of waning antibody titers (18-21). Variants such as Omicron BA.1 appear to cause less severe disease in vaccinated individuals due to cross-reactivity between the vaccine epitopes and those in the BA.1 variant, but this protection is not afforded against all Omicron variants including BA.4 and BA.5 (22-24). Whether new vaccine formulations or vaccination schemes are required to maintain lasting protection is currently an area of interest (25).

In infected individuals, the half-lives of IgG anti-spike and anti-RBD have been reported to be 103-126 and 83-116 days, respectively (26, 27). The half-life of antibodies in vaccinated individuals may be shorter, as titers are significantly decreased after 6 months (28-33). The difference in antibody half-life between infected and vaccinated individuals may depend on the half-lives of the plasma cells or differences in the memory B cells that produce them (34). Memory B cells don’t participate in the immediate increase in antibody production after re-exposure to virus or vaccine, but within several days provide high levels of protective antibodies pursuant to their peri-infection conversion to plasma cells (35). The importance of memory B cells in lasting immunity to SARS-CoV-2 infection after vaccination is highlighted by findings showing that spike protein receptor binding domain (RBD) specific memory B cells survive even after anti-RBD antibodies are absent from serum (33, 36).

In addition to humoral immunity, SARS-CoV-2-specific T cells provide protection against the virus and may be particularly relevant in the case of SARS-CoV-2 variants of concern such as B.1.617.2 delta and B.1.1.529 omicron which display mutated spike proteins that can more effectively evade neutralizing antibodies (32, 37-41). The ability of the virus to escape antibody but not T cell immunity stems from the nature of the different antigenic targets on the spike protein recognized by B cells (proteins) and T cells (peptides) (7, 40, 42-45). Underlying their potential importance, the relative expansion of SARS-CoV-2 specific CD4+ and CD8+ T cells associates with COVID-19 disease severity, and T cell memory appears more durable than serum antibody titers (26, 33, 43, 46, 47). Circulating CD4+ follicular T helper cells (cTfh) are also found in the memory T cell pool. While SARS-CoV-2-specific Tfh cell are less durable than other memory T cell subsets after vaccination and may not be required for the generation of antibodies against the virus, these cells are probably important in orchestrating a productive T and B cell response to SARS-CoV-2 infection (33, 42, 48-52).

Although we have gained significant understanding about natural immunity and response to SARS-CoV-2 infection and vaccination, informative data were not generated in chronic lung disease patients, who are at highest risk of mortality and morbidity due to COVID-19 (53). Patients with lung diseases may suffer more than healthy subjects from SARS-CoV-2 infections because of underlying pulmonary limitation and/or abnormal lung immune function. Immunosuppressant drugs taken by patients with chronic lung disease can also reduce their immune responses to the SARS-CoV-2 vaccine as reported in other disease contexts (54-58). Indeed, certain conditions and treatments may significantly reduce the ability of the patient to produce anti-SARS-CoV-2 antibody (59-66).

Individuals with chronic lung disease that fail to mount an immune response to the vaccines may be unaware of their higher risk for potentially severe “breakthrough COVID” that results from new SARS-CoV-2 variants that evade antibody neutralization. This is of particular concern as masking and social distancing have been lifted in many localities. Therefore, it is critical to understand the vaccine response in high-risk chronic lung disease patients to help identify subsets of individuals who may be at greatest risk of poor outcomes. Although the greatest at-risk patients are likely those that fail to respond appropriately to the SARS-CoV-2 vaccination, simple measurement of antibodies against the RBD does not account for heterogeneity in protective immune responses to vaccination. Therefore, to reveal whether limitations in vaccine responsiveness exist within chronic lung disease patients and to better understand the heterogeneity of responses across different chronic lung diseases, we performed deep phenotyping of the humoral and cell mediated immune response to SARS-CoV-2 vaccination in a select, investigative cohort of patients with interstitial lung disease (ILD), chronic obstructive pulmonary disease (COPD), and asthma, compared to healthy subjects.

## Results

### A subset of patients with chronic lung disease exhibit reduced serum antibody titers after mRNA vaccination against SARS-CoV-2

Serum samples were used to assess SARS-CoV-2 Pfizer-BioNTech BNT162b2 and Moderna mRNA-1273 vaccine responsiveness in a cohort of 9 asthma, 8 COPD, and 15 ILD patients and 31 healthy controls (Table 1). To investigate the humoral response, we performed an in-house quantitative ELISA for serum spike RBD-specific antibodies. Serum collected between 14-231 days after the last vaccination/boost was analyzed (Fig. 1A). Asthma (p<0.35) and COPD (p<0.022) patients showed significantly reduced antibody titers 3-4 months after vaccination compared to healthy controls. 40% (6/15) of ILD patients also exhibited reduced antibody titers compared to healthy subjects. To validate these findings, serum titers from the in-house anti-RBD assay and QuantiVac ELISA (semiquantitative Spike protein IgG) were compared. As expected, samples with the highest serum anti-RBD titers, including 100% of healthy controls, were most prominent in the highest anti-spike titer bin (>350 binding antibody units (BAU)/mL) while those showing low anti-RBD titers were enriched in the lowest bin (<150 BAU/mL) (Fig. 1B). Together, these investigative findings suggest that many patients with ILD, asthma, and COPD may not achieve or maintain the same level of humoral protection after vaccination as healthy subjects.

**Table 1:** Investigative cohort of SARS-CoV-2 vaccinated patients with chronic lung disease.

|  | **Asthma**<br>**9 subjects** | **COPD**<br>**8 subjects** | **ILD**<br>**15 subjects** | **Healthy**<br>**Controls**<br>**31**<br>**subjects** | **Total**<br>**63**<br>**subjects** |
| --- | --- | --- | --- | --- | --- |
| **Age at Sample - average (range)** | 58 (43-71) | 64 (57-73) | 62 (47-73) | 50 (25-72) | 56 (25-73) |
| **Female - number (%)** | 5 (56%) | 5 (62%) | 9 (60%) | 14 (45%) | 33 (52%) |
| **Male - number (%)** | 4 (44%) | 3 (38%) | 6 (40%) | 17 (55%) | 30 (48%) |
| **Days from last vaccination to sample - average (range)** | 117 (87-156) | 138 (112-163) | 122 (84-144) | 121 (14-231) | 123 (14-231) |
| **Immunosuppressants (%)** | 6 (66%) | 4 (50%) | 9 (60%) | 0 (0%) | 19 (30%) |
| **FEV-1 Pre-Bronch % Predicted** | 76 (49-106)<br>n=8 | 63 (27-98)<br>n=7 | 77 (33-109)<br>n=15 | n/a | n/a |
| **FVC Pre-Bronch % Predicted** | 78 (61-99)<br>n=8 | 81 (55-111)<br>n=7 | 75 (34-97)<br>n=15 | n/a | n/a |
| **FEV1/FVC Pre-Bronch % Predicted** | 94 (78-109)<br>n=8 | 75 (48-101)<br>n=7 | 101 (88-115)<br>n=15 | n/a | n/a |
| **Meets GINA 4 criteria** | 3 |  |  |  |  |
| **Meets GINA 5 criteria** | 3 |  |  |  |  |

**Figure 1:**
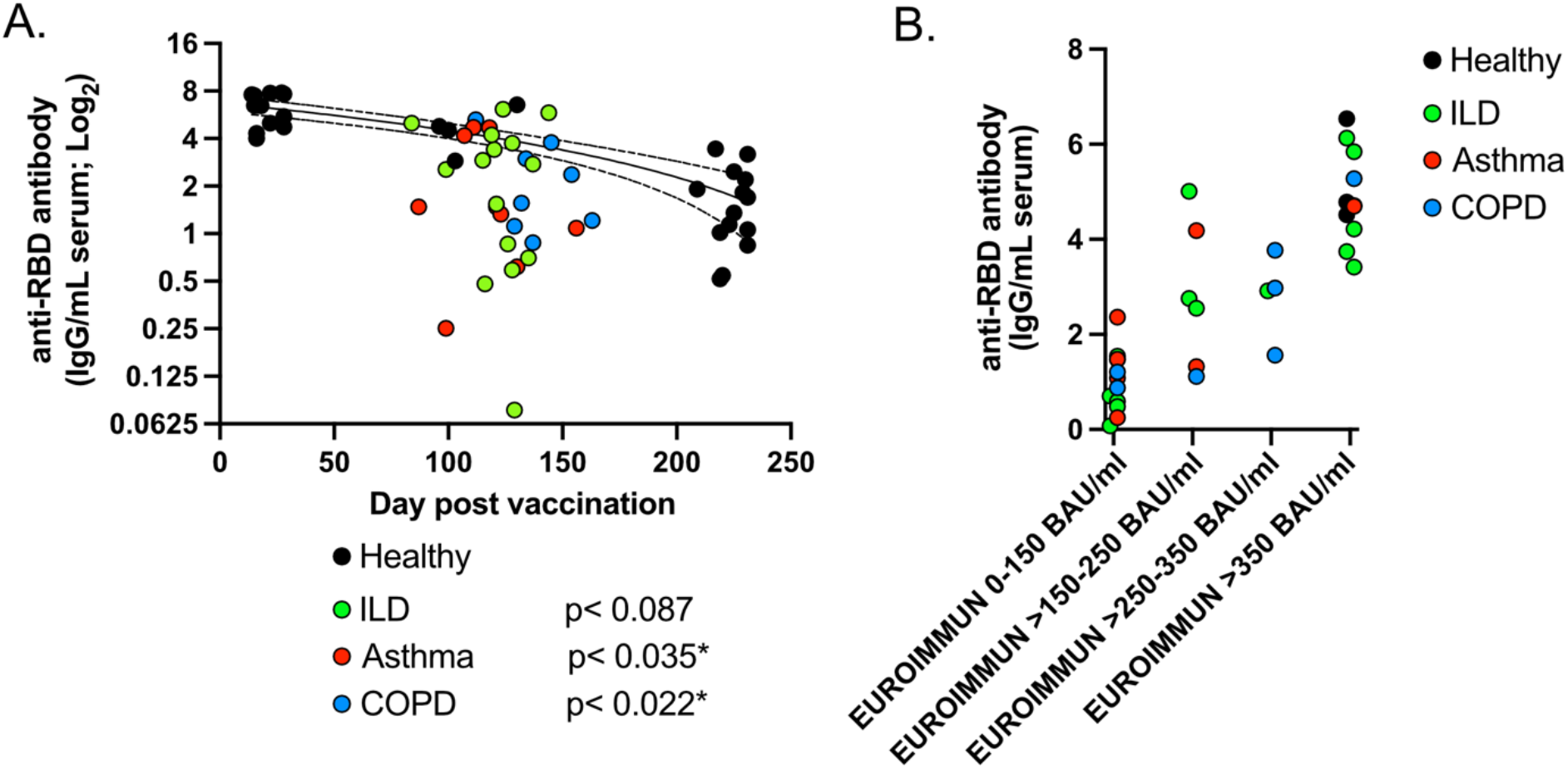
Impaired serum antibody titers against SARS-CoV-2 spike RBD in a subset of patients with chronic lung disease after vaccination. **(A)** ELISA for serum IgG binding to SARS-CoV-2 RBD in healthy (black), ILD (green), asthma (red), and COPD (blue) patients 14-231 days post SARS-CoV-2 mRNA vaccination. Line represents simple linear regression for healthy subjects flanked by 95% confidence intervals. **(B)** Serum anti-RBD antibodies detected 75-175 days after vaccination of healthy and chronic lung disease patients using the in-house ELISA were compared to antibody titers against the SARS-CoV-2 spike protein S1 domain using QuantiVac ELISA (EUROIMMUN) IgG binding antibody units (BAU). Statistical analysis: p values in represent unpaired, T test comparing titers taken 75-175 days after last vaccination.

### Circulating spike-specific B cells are reduced in patients with chronic lung disease

To investigate vaccine-specific memory B cells, we enriched PBMC for B cells and identified RBD-specific B cells using double colored RBD-tetramers (Fig. 2A) (67). We minimized contamination of non-RBD-specific B cells by eliminating B cells that bound an “irrelevant” ovalbumin-FITC protein (68, 69). Individuals with ILD (p<0.012) and asthma (p<0.032) had significantly fewer circulating RBD-specific B cells than healthy controls (Fig. 2B). COPD patients on average had fewer RBD-specific B cells within the circulating B cell population than observed in healthy controls (Fig. 2B). When RBD-specific B cells from all patients were compared to their RBD-specific serum antibody titers, a significant correlation (r=0.477; p<0.002) was observed (Fig. 2C). While the strongest positive correlation was observed in healthy subjects (r=0.572), ILD patients (r=0.588; p<0.021) also correlated. Together, these data indicate that many individuals with chronic lung disease fail to generate a robust pool of circulating vaccine-specific B cells compared to healthy controls.

**Figure 2:**
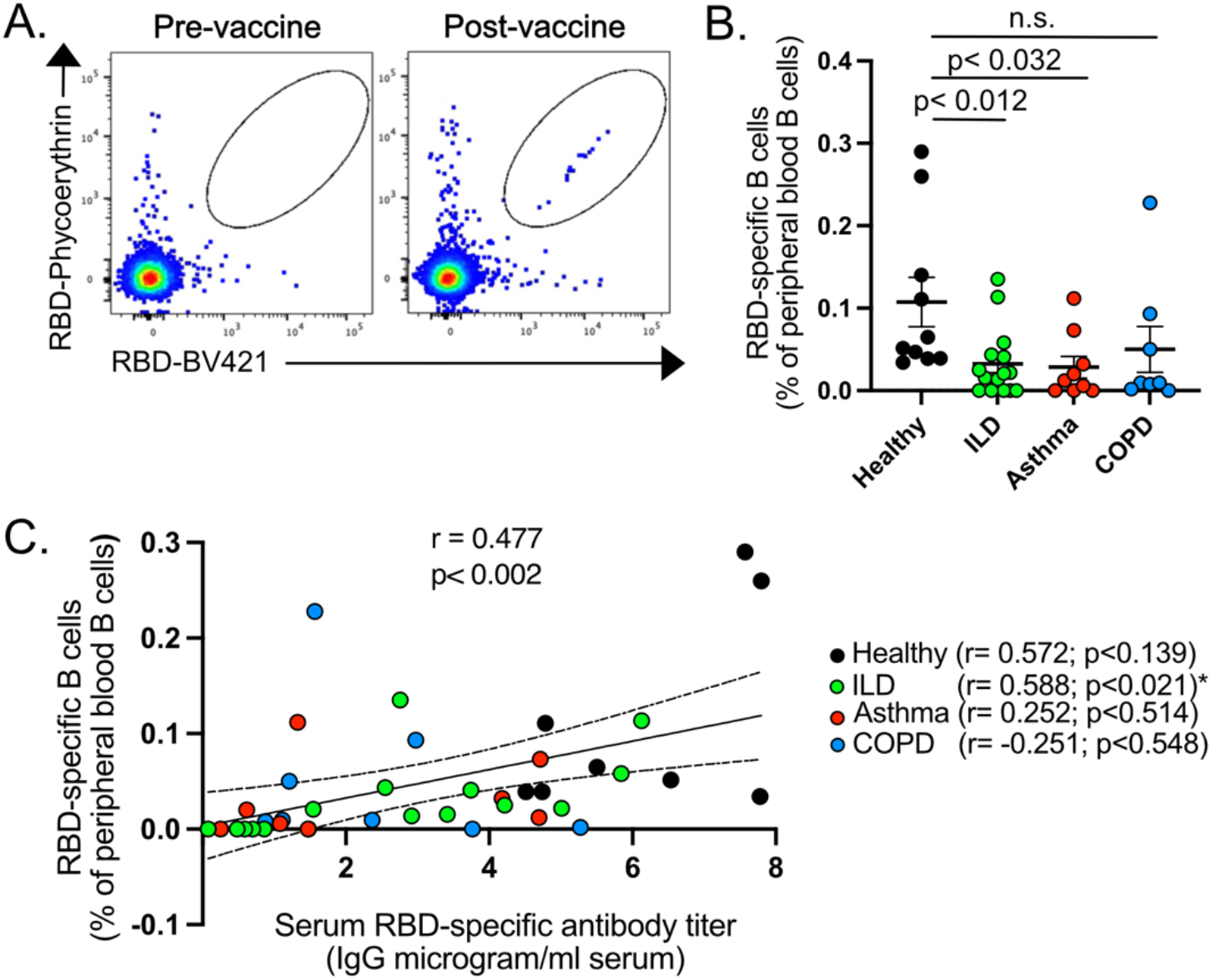
Decreased circulating RBD-reactive memory B cells in patients with chronic lung disease after vaccination compared to healthy controls. PBMC collected 14-175 days post vaccination. **(A)** Representative contour plots of circulating B cells from blood of patients pre- and post-SARS-CoV-2 vaccination. Gate represents dual RBD-tetramer binding B cells. **(B)** Graph represents the percentage of RBD+ B cells within the total circulating B cell pool of healthy (black), ILD (green), asthma (red) and COPD (blue) patients. **(C)** Correlation of serum anti-RBD antibody titers and circulating RBD-binding B cells detected in healthy and chronic lung disease patients after SARS-CoV-2 vaccination. Lines represents best-fit simple linear regression with flanking lines demarcating 95% confidence intervals. (n=8-14; error bars represent +/- S.E.M)

### T cell response to SARS-CoV-2 vaccination is impaired in patients with chronic lung disease

To investigate the RBD-specific CD8+ and CD4+ T cell responses in a way that was agnostic to a patient’s HLA type, we used a modified approach previously described to efficiently detect spike-responsive T cells in the blood of patients with mild COVID-19 (48). Using this approach, subsets of individuals with underlying lung conditions exhibited diminished RBD-specific T cell responses compared to healthy controls (Fig. 3A, B). Specifically, CD8+ (p<0.004) and CD4+ (p<0.023) T cell responses in asthma patients were significantly reduced, as were CD8+ (p<0.008) T cell responses in COPD patients. Of note, 21% of ILD patients showed limited CD8+ T cell responses and 42% failed to evoke a robust CD4+ T cell response after vaccination. Similarly, 33-37.5% of asthmatic and COPD patients had no observable CD4+ and CD8+ T cell responses to the vaccine antigen. While CD4+ T cell responsiveness correlated strongly (r=0.728; p<0.0001) with CD8+ T cell vaccine responses across disease cohorts, no correlation was observed between RBD-specific T cell responses and RBD-specific antibody titers (Fig. 3C-E). This suggests that an individual’s humoral vaccine response can be independent of their vaccine-elicited T cell immunity and vice versa.

**Figure 3:**
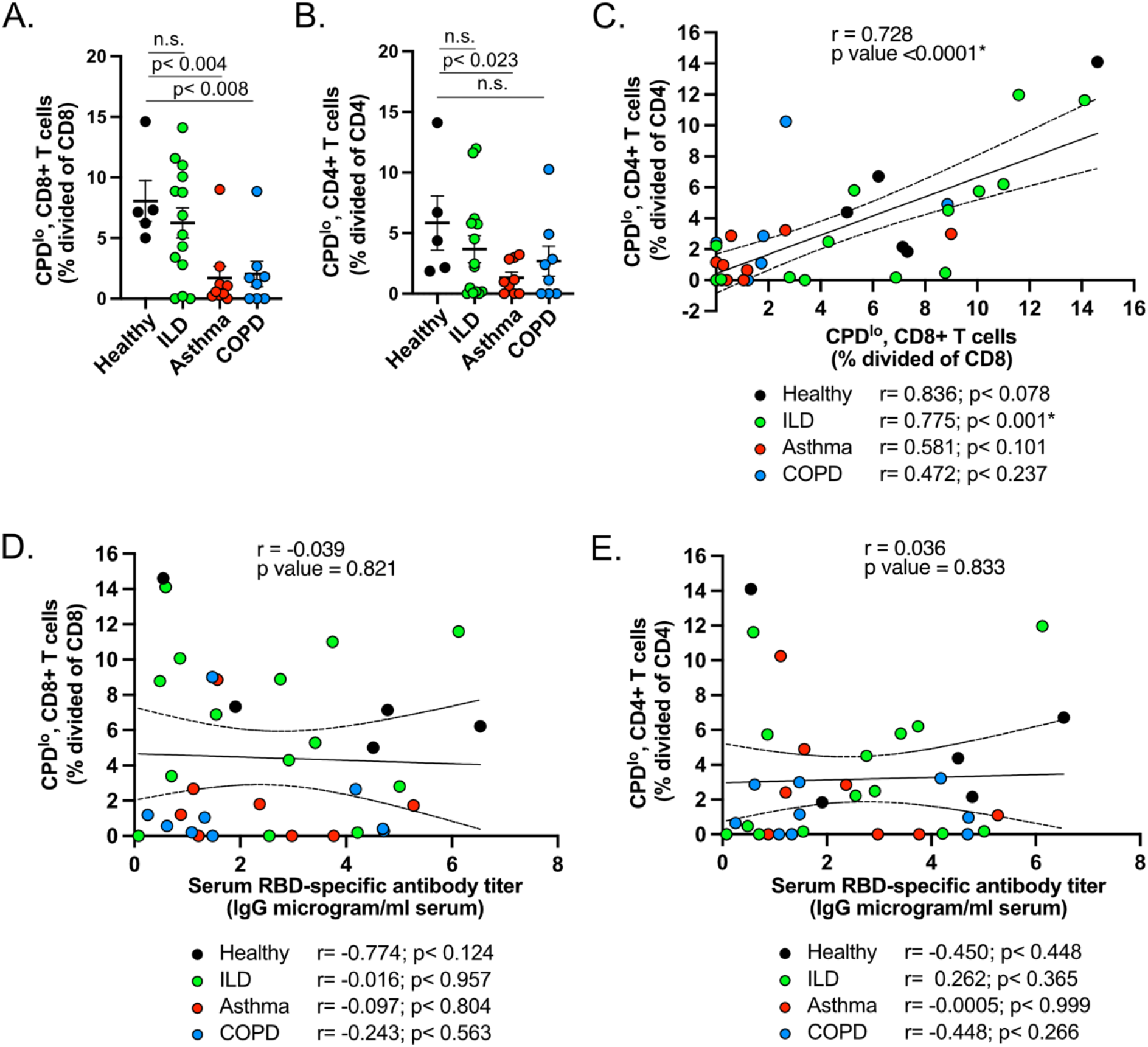
Decreased circulating SARS-CoV-2 RBD-specific T cells in a subset of patients with chronic lung disease after vaccination. PBMC collected 75-220 days post vaccination. **(A)** Graph represents the percentage of CPD-low (divided) RBD-specific CD8+ T cells within the total circulating CD8+ T cell populations after culture and stimulation with RBD-protein in healthy (black), ILD (green), asthma (red) and COPD (blue) patients. Numbers are normalized by subtracting CPD-low population in PBMC cultures that received no protein. (**B)** Graph represents the percentage of CPD-low (divided) CD4+ T cells within the total circulating CD4+ T cell populations after culture and stimulation with RBD-protein in healthy patients and patients with chronic lung disease. Numbers are normalized by subtracting CPD-low population in PBMC cultures that received no protein. **(C)** Correlation of CPD-low (divided) CD4+ and CD8+ T cells in circulation in healthy (black), ILD (green), COPD (blue), and asthma (red) patients after SARS-CoV-2 vaccination. Central line represents best-fit simple linear regression; flanking lines demarcate 95% confidence intervals. **(D)** Correlation between CPD-low (divided) CD8+ T cells in circulation and serum antibody titers against RBD in healthy (black), ILD (green), COPD (blue), and asthma (red) patients after SARS-CoV-2 vaccination. Central solid line represents best-fit simple linear regression; flanking lines demarcate 95% confidence intervals. **(E)** Correlation between CPD-low (divided) CD4+ T cells in circulation and serum antibody titers against RBD in healthy (black), ILD (green), COPD (blue), and asthma (red) patients after SARS-CoV-2 vaccination. Central line represents best-fit simple linear regression; flanking lines demarcate 95% confidence intervals. (n=5-14; error bars represent +/- S.E.M)).

### Vaccine-specific T cells in patients with chronic lung conditions have impaired cytokine potential

To address T cell function, the cytokine potential in our patient cohorts was assayed by intracellular cytokine staining. While the percentages of bulk CD8+ T cells that were IFN-γ competent were significantly (p<0.012) elevated among vaccinated COPD patients compared to healthy controls, the percentage of such cells in asthmatic and ILD patients were not significantly different (Fig. 4A). On the other hand, the percentage of bulk CD8 T cells from asthmatic patients that could produce IL-2 were significantly (p<0.014) reduced relative to healthy controls (Fig. 4A). While this suggests some heterogeneity exits in the cytokine profiles of patients with chronic lung disease, for the most part, bulk T cell function appears similar across disease groups. Even less heterogeneity was observed in the cytokine potential of CD4+ T cells across disease groups and healthy patients (Fig. 4B).

**Figure 4:**
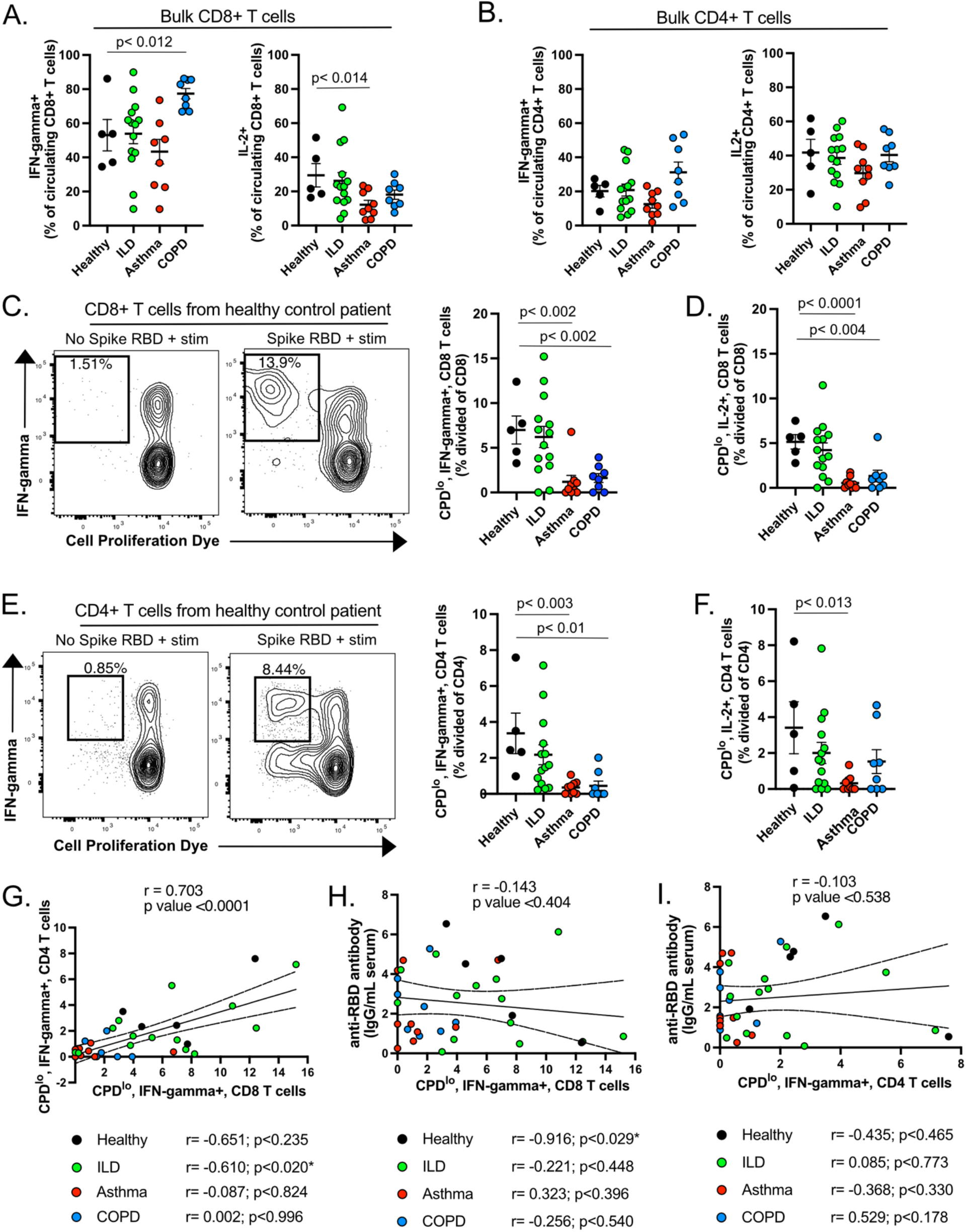
Impaired cytokine potential among SARS-CoV-2 RBD-specific T cells after vaccination of patients with chronic lung disease. PBMC collected 75-220 days post vaccination. **(A)** Graph represents the percentage of IFN-gamma and IL-2 expressing CD8+ T cells within the total circulating CD8+ T cell population after stimulation with RBD-protein in healthy patients and patients with chronic lung disease. **(B)** Graph represents the percentage of IFN-gamma and IL-2 expressing CD8+ T cells within the total circulating CD8+ T cell population after stimulation with RBD-protein in healthy patients and patients with chronic lung disease. **(C)** Contour plots and graph identifying the percentage of IFN-gamma+ CPD-low (divided) CD8+ T cells within the total CD8+ T cells pool. Gate in contour plot identifies circulating CPD-low (divided) CD8+ T cells that express IFN-gamma. Dividing cells above background are only found in cultures stimulated with RBD. **(D)** Graph identifying the percentage of IL-2+ CPD-low (divided) CD8+ T cells within the total CD8+ T cells pool. Numbers in graph are normalized by subtracting CPD-low (dividing) population in PBMC cultures that received no protein. **(E)** Contour plots and graph identifying the percentage of IFN-gamma+ CPD-low (divided) CD4+ T cells within the total CD4+ T cells pool after vaccination. Gate in contour plot identifies circulating CPD-low (divided) CD4+ T cells that express IFN-gamma. Notable CPD^low^ population that falls outside of gate represents RBD-responsive T cells that are not expressing IFN-gamma. Numbers in graph are normalized by subtracting CPD-low (dividing) population in PBMC cultures that received no protein. **(F)** Graph identifying the percentage of IL-2+ CPD-low (divided) CD4+ T cells within the total CD4+ T cells pool. Numbers in graph are normalized by subtracting CPD-low (dividing) population in PBMC cultures that received no protein. **(G, H, I)** Correlation between IFN-gamma expressing CPD-low (divided) CD4+ and CD8+ T cells (G), IFN-gamma expressing CPD-low (divided) CD8+ T cells and serum RBD antibody titers (H) and IFN-γ expressing CPD-low (divided) CD4+ T cells and serum RBD antibody titers (I) in healthy (black), ILD (green), asthma (red), and COPD (blue) patients after SARS-CoV-2 vaccination. Central solid line represents best-fit simple linear regression; flanking lines demarcate 95% confidence intervals. (n=5-14; error bars represent +/- S.E.M)).

In contrast to bulk T cell populations, heterogeneity in cytokine potential was observed in vaccine-responsive T cell populations. In these experiments vaccine responsive T cells were defined by loss of CPD, indicative of cells that had divided in response to RBD antigen. In patients with chronic lung disease, the percentage of RBD responsive CD8+ T cells from asthma and COPD patients that could produce IFN-γ and/or IL-2 was significantly reduced compared to similar T cells obtained from healthy subjects (Fig. 4C, D). A similar finding was observed in RBD responsive CD4+ T cells from asthma and COPD patients (Fig. 4E, F). Of note, while asthma and COPD patients showed more homogeneity in their T cell functionality, a subset of patients with ILD also exhibited decreased IFN-gamma and IL-2 within RBD-specific CD4+ and CD8+ T cells compared to healthy controls. This suggests that at least some patients within each disease cohort exhibit reduced T cell functionality to the vaccine.

When looking at total T cell responsiveness, patients mounting a productive CD4+ T cell response generally exhibited a productive CD8+ T cell response (r=0.703; p<0.0001) (Fig. 4G). To understand if T cell function similarly tracked with humoral immunity after vaccination, we compared IFN-gamma+ RBD-specific T cells in each patient to their serum anti-RBD titers. In all patient groups, no significant correlation was observed (Fig. 4H, I). Together with serum antibody and memory B cell data, these findings indicate that the SARS-CoV-2 vaccine may differentially promote T cell and humoral immunity in some ILD, asthma, and COPD patients.

### SARS-CoV-2-specific Tfh cells exhibit decreased cytokine potential in patients with chronic lung conditions compared to healthy controls

Given Tfh cells are important in driving humoral vaccine responses, we next investigated the Tfh response in vaccinated patients with pulmonary disease. The percentage of circulating CXCR5+ CD4+ Tfh (cTfh) cells among the total CD4+ T cell pool was decreased across all disease cohorts reaching significance within asthma (p<0.011) and COPD (p<0.006) patients (Fig. 5A). While IL-2 production remained comparable to healthy controls, the relative percentage of IFN-γ expressing cTfh cells was increased across all chronic lung disease cohorts (Fig. 5B). Increased IFN-γ production was most evident in COPD (p<0.027) patients, however, at least some ILD and asthma patients also exhibited increased interferon expression within bulk cTfh cells relative to healthy controls. Despite the increased IFN-γ production observed in bulk cTfh cells in patients with chronic lung disease, RBD-responsive (CPD-lo) cTfh on average exhibited decreased IFN-γ production compared to vaccinated, healthy controls. In fact, 21% of ILD patients, 44% of asthma patients, and 25% of COPD patients in this investigative cohort lacked IFN-γ-expressing RBD-responsive Tfh cells above background (Fig. 5C). This mirrors the decreased functionality of vaccine responsive T cells within non-Tfh cell populations.

**Figure 5:**
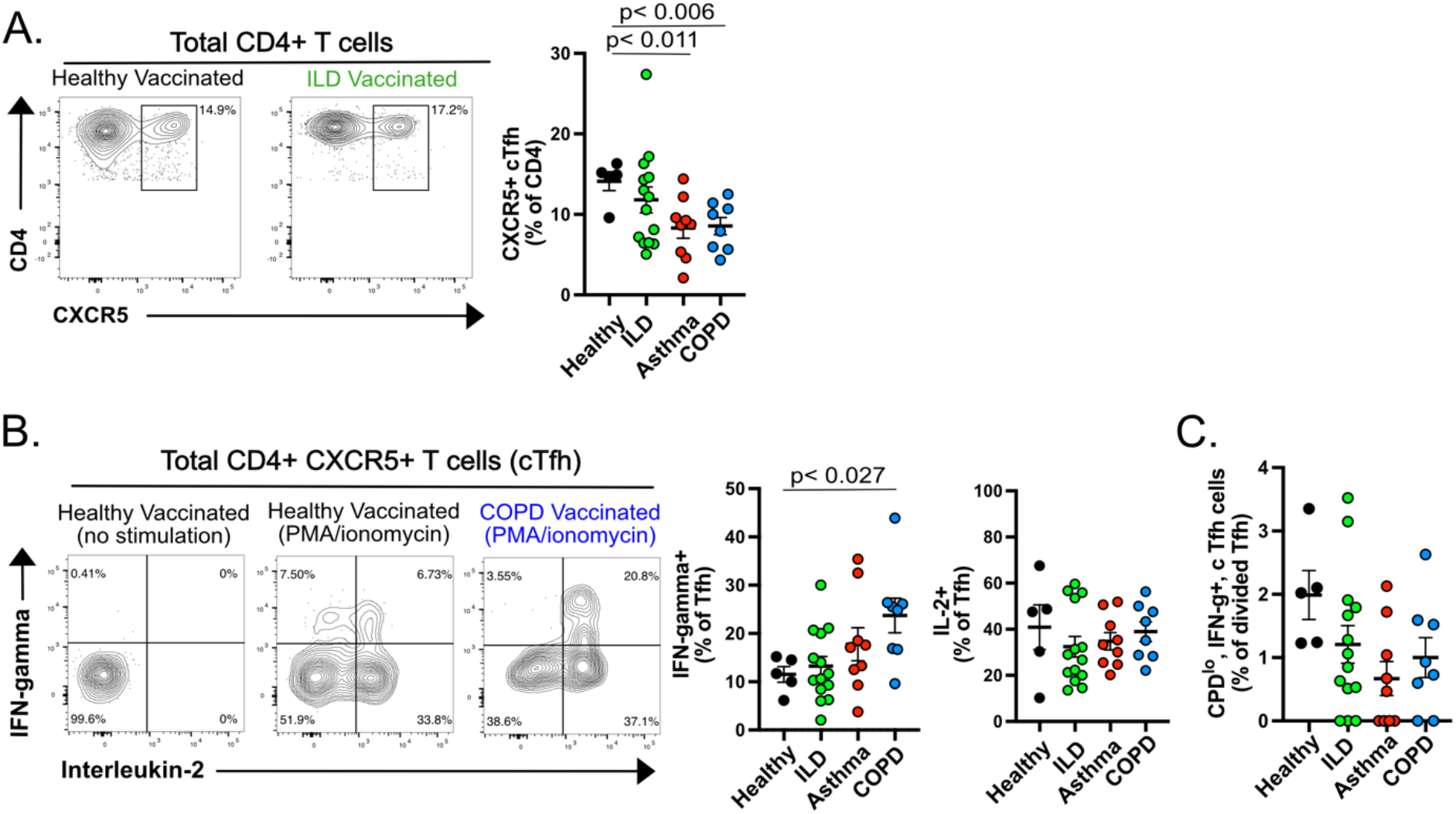
Patients with chronic lung disease have heterogeneous Tfh cell responses after SARS-CoV-2 vaccination compared to healthy controls. PBMC collected 75-220 days post vaccination. **(A)** Contour plots from representative PBMC cultures from healthy and ILD vaccinated patients. Gate reveals the percentage of CXCR5^+^ Tfh cells among total circulating CD4^+^ T cells. **(B)** Representative contour plots of CXCR5+ Tfh cells from PBMC cultures of healthy and COPD vaccinated patients with or without stimulation with PMA/ionomycin. Gates reveal the percentage of CXCR5^+^ Tfh cells expressing one of or both IFN-γ and IL-2 cytokines. Graphs show the percentage of cTfh in these distinct disease cohorts and healthy controls that express IFN-γ or IL-2. **(C)** Graph shows the percentage of IFN-γ expressing CPD-low (divided) CXCR5+ Tfh cells within the RBD-specific Tfh cell population in healthy patients and patients with chronic lung conditions. Numbers in graph are normalized by subtracting CPD-low (dividing) population in PBMC cultures that received no protein. (n=5-14; error bars represent +/- S.E.M).

## Discussion

This study highlights the significant heterogeneity that exists in the vaccine response to SARS-CoV-2 in individuals with ILD, COPD and asthma compared to healthy controls. In our assessment of vaccine-induced antibody titers, memory B cell subsets, and T cells in patients with asthma, COPD, and ILD, we found that 48.3% of patients with chronic lung disease exhibited serum antibody titers to the vaccine antigen below the expected titers observed in healthy controls 3-4 months after the last vaccine administration. This correlated with decreased RBD-specific circulating memory B cells. In addition to impaired humoral hallmarks, most patients with asthma and COPD and a subset of patients with ILD had reduced circulating RBD-responsive CD4+ T cells, CD8+ T cells, and Tfh cells. These vaccine-specific T cell populations also exhibited decreased cytokine potential compared to healthy controls. Of note, while some individuals lacking antibody and memory B cell production after vaccination also exhibited reduced T cell immunity, many patients had evidence of defects in only one arm of the adaptive response to SARS-CoV-2 vaccination. This highlights the considerable variability in vaccine responses among patients with chronic lung disease and illustrates the importance of deep immunophenotyping of high-risk patients to determine their overall immunity to SARS-CoV-2 after vaccination.

Most of the available data regarding the safety, efficacy, and durability of mRNA vaccines against SARS-CoV-2 has been generated from healthy vaccinated cohorts (5, 20, 29, 33, 70-73). In these initial studies, nearly all healthy vaccine recipients developed binding and neutralizing antibodies. However, this level of vaccine responsiveness does not appear to always extend to individuals with chronic lung disease (59, 74). While not designed or powered to address safety, efficacy, or durability of the vaccine response in patients with chronic lung disease, the current data suggest that what we understand regarding vaccination in healthy subjects may not be directly applicable to patients with chronic lung disease. Further, the data also show that vaccine responses may differ depending on the type of underlying lung condition. For example, as a group, individuals with COPD and asthma were more likely to exhibit impaired antibody and T cell responses than ILD patients, who instead exhibited greater heterogeneity in their mRNA vaccine response. Factors that separate responders from non-responders within a particular disease group may reflect distinct disease-associated endotypes within COPD, asthma, and ILD, including the possibility that subsets of each of these lung diseases are associated with broadly abnormal immunity, a concept that finds support in previous studies (75). Understanding how the intrinsic nature of each pulmonary disease impacts B cell and T cell immunity in patients with chronic lung disease is particularly important as such patients are more at risk for “breakthrough COVID-19” driven by emerging SARS-CoV-2 variants of concern.

One of the key caveats in the current study is the lack of a longitudinal assessment within these different disease cohorts. We know from healthy controls that each arm of the immune system varies over time after vaccination. For example, while anti-RBD antibody titers and cTfh numbers wane six months after vaccination, vaccine-specific T cell responses and memory B cell responses remain relatively stable over that same period in healthy subjects (33, 76). Whether similar kinetics occur in individuals with chronic lung disease remains unknown. The investigative data provided herein suggest that a large percentage of individuals with chronic lung disease fail to mount productive humoral and cell mediated immunity during the first and second dosing of the vaccine. What remains unclear is whether such non-responders remain impaired after subsequent vaccination attempts. While there is evidence that a third booster can be effective in providing some protection against SARS-CoV-2 in other high-risk populations (62, 77-79), some seronegative individuals who did not respond to the first two doses of vaccine also fail to respond to the third boost (80). How boosting can benefit non-responders becomes even more complicated as natural exposures to the virus and its variants become more frequent. Thus, the benefit of multiple boosts or more frequent boosting in subsets of patients with asthma, ILD, and COPD that show inadequate vaccine responsiveness should be explored.

In conclusion, vaccination against SARS-CoV-2 has had a significant impact on our ability to control the current COVID-19 pandemic. However, much of what we understand comes from data collected from clinical trials comprised of healthy individuals. Our study suggests that efficacy of the vaccine and vaccine-induced immunity in healthy individuals should not be uniformly extrapolated to individuals with chronic lung disease. This finding has clinical relevance, as these individuals are considered at high-risk for contracting severe COVID-19. Patients with COPD, for example, have increased odds of hospitalization, intensive care unit admission, and mortality compared to healthy controls if exposed to SARS-CoV-2 (53). Given the relatively high percentage of patients with chronic lung disease showing some form of impaired vaccine responsiveness and the high degree of heterogeneity in the responses observed across individuals with ILD, asthma, and COPD, chronic lung disease patients may benefit from personalized vaccination schemes and deeper assessment of immune responses to ensure optimal protection in this vulnerable population.

## Methods

### Study participants

Chronic lung disease and healthy control blood samples were collected as part of two institutional IRB-approved protocols under which subjects provided informed consent: 1) a prospective study of response to SARS-CoV-2 vaccinations that recruited from NJH clinics and 2) the National Jewish Health BioBank that recruits patients undergoing normal clinical laboratory testing or from a healthy donor pool. The samples were stored and maintained as part of the National Jewish Health (NJH) Biobank. Patient information regarding vaccine status, medicine, and infection status was collected at time of sample collection or as part of their normal medical record.

### Serum and peripheral blood mononuclear cell sample preparation

Blood was collected from multiple 10 mL blood draws into EDTA tubes. Serum was processed after density gradient centrifugation and PBMC –post red blood cell lysis– were resuspended in 10% DMSO + 90% FBS in cryovials prior to storage in liquid nitrogen.

### SARS-CoV-2 receptor binding domain generation

SARS-CoV-2 spike receptor binding domain (aa319 to aa541) with C-terminal 6* histidine tag was expressed in 293F cell as described previously (81). The RBD protein was purified with nickel column and the eluted protein was further purified by size-exclusion column to collect monomer sized RBD.

### RBD-tetramer generation

SARS-CoV-2 spike receptor binding domain (aa319 to aa541) with C-terminal histidine tag and Avitag was expressed and purified in the same way above. The RBD was biotinylated by BirA enzyme. The biotinylated RBD was conjugated to the streptavidin labeled with different fluorescent dyes.

### Enzyme-linked immunosorbent assay (ELISA) for RBD serum antibody

Twenty µg/ml 6*-histidine tagged RBD was used for coating ELISA plate. After blocking, human serum at different dilutions was incubated on the plates. The bound IgG was detected with goat anti-human IgG, Fcγ fragment specific conjugated with alkaline phosphatase (Jackson ImmunoResearch #109-055-008). Bamlanivimab was used as standard for converting ELISA O.D. value to serum antibody amount (82).

### Staining of RBD specific B cell subsets by flow cytometry

Human PBMC samples were obtained from Biobank at National Jewish Health. Cells were stained with 2 µg/ml double colored RBD tetramers (conjugated with BV421 and PE respectively), human Fc block and FITC-OVA first on ice for 30 minutes. CD19 APCcy7, IgD BV510, dump (CD4, CD8, CD14, CD16) PerCP antibodies were then added for staining. Cells were washed and stained with Ghost UV450 dye and fixed with 1% paraformaldehyde for flow cytometry analysis.

### PBMC cultures and antigen-specific T cell stimulation

PBMCs were thawed and resuspended in complete RPMI-1640 (10% FBS, 10mM HEPES, 50uM 2-beta mercaptoethanol, 2mM L-glutamine, and 1% penicillin and streptomycin). After counting, PBMC were stained with 5uM cell proliferation dye eFluor 670 (CPD; #65-0840, Thermo Fisher). CPD labeled cells were plated at 2×10^5^ PBMC per well in cRPMI + 2ng/mL (10U/mL) of recombinant human IL-2 (Biolegend). For RBD stimulation, wells were incubated with 2.5ug/mL of RBD or media alone. For cytokine analysis, cultures were left unstimulated or were stimulated with 50ng/mL phorbol 12-mryistate 13-acetate (PMA; Sigma) and 1ug/mL of ionomycin (Sigma-Aldrich) 4 hours before harvest. All wells were provided 10ug/mL of brefeldin A (Sigma-Aldrich) and 1x dilution of monensin (GolgiStop; #554724 Becton Dickinson) to prevent cytokine secretion.

### Staining of T cell subsets by flow cytometry

PBMC were labeled with LIVE/DEAD fixable violet dye (L34955; Invitrogen), followed by surface antibody staining (CD4, Clone:RPA-T4; CD8, Clone:SK1; CD3, Clone:OKT3, CXCR5, clone:J252D4, Biolegend). After surface staining, cells were fixed and permeabilized using FOXP3/Transcription factor staining buffer set (#00-5523-00, Invitrogen) per manufacturer’s instructions. Fixed cells were stained for intracellular cytokines anti-IL-2 (Clone:MQ1-17412, Biolegend) and anti-interferon gamma (Clone:4S.B3, Biolegend). Data were collected by flow cytometric analysis on a LSR II (BD Biosciences) cytometer and analyzed using FlowJo (BD Bioscience).

### Statistical Analysis

All comparisons were made using paired and unpaired t tests with Prism 9 (GraphPad). Where possible p values and r correlations are provided directly in figures. P values in grouped graphs represent unpaired, two-tailed T test.

## Data Availability

All data produced in the present study are contained in the manuscript or are available upon reasonable request to the authors

## Acknowledgments

We thank John Yang management and distribution of PBMC and serum samples from NJH Biobank and the Jin Hua foundation for their support.

## Author contributions

Concept and design: all authors. Acquisition and interpretation of data: H.L., K.A., P.Z., S-Y. L., P.M., R.L.R. Drafting of the manuscript: H.L., P.M., R.L.R. Critical revision of the manuscript for important intellectual content: All authors. Statistical analysis: R.L.R., H.L., S.-Y.L., P.Z. All authors had full access to all the data in the study and take responsibility for the integrity of the data and accuracy of the data analysis.

